# Exploring intersectional determinants of, and interventions for, low uptake of human papillomavirus vaccine in Sub-Saharan Africa: A scoping review protocol

**DOI:** 10.1101/2024.10.11.24315319

**Authors:** Peter N Kailemia, Victoria N Mukami

**Author notes:** **Authors twitter/X handle:** @kailemia1; @NdegwaViki.

## Abstract

**Introduction:** Cervical cancer is the most diagnosed cancer and the leading cause of cancer death in 36 low and middle-income countries with the majority being located in sub-Saharan Africa (SSA), South America, and South Eastern Asia. The highest regional incidence and mortality occur in SSA. Despite the high efficacy and cost-effectiveness of the HPV vaccine in preventing cervical cancer, its uptake remains unacceptably low in SSA. This scoping review aims to integrate evidence from SSA on social determinants of HPV vaccine uptake with complementary evidence on interventions to promote its uptake.

**Methods and analysis:** The proposed review will be conducted following the guidelines by the Joanna Briggs Institute Scoping Review Methodology Group. Additionally, sequential explanatory design will guide the integration of *determinants evidence* with *interventions evidence*. This scoping review will be reported per the Preferred Reporting Items for Systematic Reviews and Meta-Analysis Extension for Scoping Reviews (PRISMA-ScR) checklist. Five databases, PubMed/MEDLINE, LIVIVO, Google Scholar, BASE (Grey Literature), Preprints databases (e.g. OSF and MedRxiv), and African Journals Online (AJOL) will be searched, with results limited to English language publications and those published from 2006 to 2024. Two forms will be used for data extraction from the determinants and interventions studies by two independent reviewers. A narrative summary of evidence from the both determinants and interventions studies will be conducted. Furthermore, a multi-level analysis will be conducted to explore the intersections of determinants across socioecological levels of health behaviour. A further integrative cross-study analysis of results from determinants and interventions studies will be conducted where the determinants evidence will be used to interrogate the intervention evidence. Data will be presented in tables and matrices.

**Ethics and dissemination:** No ethical approval will be required for this study because it will be based on data collected from publicly available records. The review results will be disseminated widely through a peer-reviewed publication and other forums such as workshops, conferences, and meetings with local health administrators, policymakers and other wider stakeholder engagements.This protocol has been registered with Open Science Framework (https://doi.org/10.17605/OSF.IO/5JKZ8)

## INTRODUCTION

Cervical cancer is the fourth most frequently diagnosed cancer with an estimated 604,000 new cases and the fourth leading cancer of mortality with 342,000 deaths worldwide in 2020 ^1^. Additionally, Cervical cancer is the most diagnosed cancer and cause of cancer death in 36 low-and middle-income countries (LMIC) with a majority being located in sub-Saharan

Africa (SSA), Melanesia, South America and South Eastern Asia^1^. The highest regional incidence and mortality occur in SSA, particularly in Eastern Africa, Southern Africa, and Middle Africa^1^. Conversely, in high-income countries (HICs) such as Unites States, Australia, and New Zealand, the incidence rate and mortality rates are much lower at approximately 8 and 18 times lower respectively^2^.

Although there are many risk factors for cervical cancer such as HIV, smoking, Chlamydia Trichomatis, higher number of childbirths and long-term use of oral contraceptives, Human Papillomavirus (HPV) is the main etiological factor^3,4^. HPV prevalence in SSA is among the highest at an estimated average of 24%^5^. Compelling evidence suggests that populations in lower socioeconomic settings have a greater risk of exposure to risk factors for cervical cancer^6^. Lower socioeconomic status and increased exposure to HPV largely explain the high prevalence and mortality rates in LMICs, including SSA^7,8^.

Primary prevention measures (HPV vaccine) and secondary ones (screening) are highly effective in the prevention and early detection of cervical cancer respectively^1^. However, there are wide disparities in the implementation of these measures between LMICs and HICs. Studies suggest that while >60% of women from HICs have ever been screened for cervical cancer, only rates as low as 16.9% have been achieved in most countries in SSA^8^. While several factors may explain the low screening rates in SSA, it is reasonable to argue that the limited resources in these settings are a major barrier to the establishment of population-based screening programs. Evidence suggests that the HPV vaccination reduces the burden of cervical cancer by 90% ^9^. Currently, the World Health Organisation (WHO) recommends a 2-dose HPV vaccine for girls 9 to 13 years as the most efficacious and cost-effective intervention for long-term reduction in cervical cancer burden^10,11^. In light of this, WHO in 2020, set an ambitious global strategy of ensuring 90% of girls are fully vaccinated by the age of 15 years ^12^.

Despite the HPV having been introduced since 2006, as of 2020 only 22 of the 78 lower and lower-middle-income countries had introduced the vaccine compared to 35 of 59 upper-middle-income countries and 50 of 57 HIC^13^. Consequently, only 25% of adolescents living in lower and lower-middle-income countries have access to the HPV vaccine^13^. Consistent with other LMICs, the HPV vaccine uptake remains low in SSA^8,14,15^. A recent systematic review on HPV vaccine uptake in SSA has identified various determinants such as the healthcare system, socioeconomic status, stigma, experience with vaccines, health education, policy, stakeholder engagement, and women’s empowerment^7^ as drivers of the vaccine uptake.

Considering the low uptake of HPV vaccine in SSA and other parts of the world, there have been attempts to develop and implement interventions to promote uptake. Most of the current interventions implemented in SSA, however, are single-level educational interventions with limited effectiveness ^16-18^. Notably, the interventions lack multilevel and intersectional focus despite strong evidence showing that social determinants of health behaviour occur at multiple levels^19^ and intersect both within and across these levels^20,21^. Furthermore, a recent systematic review suggests that adopting an intersectional lens in cancer care has the potential to promote multidimensional and holistic care across the cancer continuum ^22^

While there is evidence on the social determinants of HPV vaccine uptake in SSA, and interventions have been implemented to promote vaccine uptake, the uptake and adherence remain low^8^. Previous reviews on social determinants and interventions of HPV vaccine uptake^7^ have ignored intersectional interactions of social determinants within and across socio-ecological levels of health behaviour. Furthermore, they have considered evidence on the uptake of the HPV vaccine from a *siloed* perspective, where they have exclusively focused on determinants ^7,23,24^ of, or interventions^16-18^ for the promotion of the vaccine uptake. The persisting low uptake raises questions about the extent of alignment between existing interventions and social determinants of vaccine uptake.

The authors of this protocol use the term *contextual determinants-sensitivity* of behaviour change interventions to bring to the fore the importance of ensuring interventions are sensitive to the contextual drivers of the target behaviour within a particular population. Evidence from the field of health psychology^25^, various intervention development frameworks^26,27^, and other literature^28^ strongly suggests that considerations of contextual determinants of behaviour ensures that interventions are culturally-sensitive. For instance, among the Arabic-speaking immigrant population in Australia, lack of access to the Arabic language version of HPV vaccine educational materials as well as religious factors were identified as uniquely important contextual determinants of vaccine uptake^29^. It is not known if the current interventions to promote HPV vaccine uptake are aligned with intersectional determinants of vaccine uptake in SSA. Ensuring that interventions for promoting vaccine uptake are aligned with the contextual drivers of low uptake in SSA will progress the region towards the WHO goal of 90% vaccination levels^12^.

The current review attempts to narrow this gap by integrating evidence on social determinants of HPV vaccine uptake with complementary evidence on interventions for the promotion of its uptake in SSA. Furthermore, this study attempts to narrow a methodological gap identified in previous reviews around the integration of evidence on behaviour change interventions with evidence on target behavioural determinants^30-32^.

A preliminary search of Google Scholar, Google, Open Science Framework, and JBI Evidence Synthesis database was conducted between October and November 2023 to determine if scoping reviews or other reviews using the methods proposed in this protocol were published or ongoing. The search identified *siloed (*isolated) reviews examining determinants of vaccine uptake as well as reviews on interventions to promote its uptake. To judge the degree of *contextual determinants-sensitivity* of interventions, no scoping review or any other type of review attempted to integrate HPV vaccine uptake *determinants evidence* with *interventions evidence* to promote its uptake.

While there are many indications for conducting a scoping review such as being a precursor for a systematic review, mapping out available evidence in a field, analysing knowledge gaps, clarifying key concept definitions, and examining how research is conducted in a field^33,34^, reviewers have to be explicit about the choice of such review type. Considering the *siloing* of the current evidence around HPV vaccine uptake among girls, this scoping review aims to identify and analyse gaps in the integration of *determinants evidence* with *intervention evidence* about vaccine uptake in SSA. Furthermore, the review attempts to map the types of evidence available on this topic. The evidence produced from this review may stimulate further evidence synthesis efforts on the topic. To the best of the authors’ knowledge, this is the first scoping review on HPV vaccine uptake to attempt to uniquely integrate *determinants evidence* and *interventions evidence* to inform efforts around HPV vaccine uptake.The review protocol has been registered in the Open Science Framework (DOI: https://doi.org/10.17605/OSF.IO/5JKZ8)

### Review aims and questions

This scoping review aims to integrate evidence from SSA on social determinants of HPV vaccine uptake with complementary evidence on interventions to promote its uptake. The review will be guided by the following questions:

- What are the barriers to, and facilitators for the uptake of the human papillomavirus vaccine among the youth in SSA?
- What is the effectiveness of interventions for the promotion of human papillomavirus vaccine uptake among adolescents in SSA?
- What interventions address the reported barriers to HPV vaccine uptake or build upon facilitators to promote its uptake in SSA?

### Eligibility criteria

The construction of the eligibility criteria was guided by the PCC (population, concept, and context) Framework^35,36^. Although the general purpose of scoping reviews is to provide a map of available evidence rather than synthesise evidence for informing policy and practice^33^, the purpose and nature of the review questions influence the specific eligibility criteria of included studies^35^.To this end, the PCC framework will be flexibly applied considering the focus of the review questions.

### Participants/population

Participants will include adolescent girls aged between 9 and 19 years, the age at which the vaccine is most effective ^1^. Studies that target parents/caregivers will also be included as they indirectly influence the healthcare decisions of their children especially those below 18 years, the legal decision-making age in most countries. Studies that involve other populations either separately or in combination with adolescent girls and/or parents will be excluded.

### Concept

Records will be considered for inclusion in the review if they focus on social determinants that may have directly or indirectly (through parents/caregivers) influenced adolescent girls’ uptake of the HPV vaccine. Records will be considered if they have focused on barriers to, or/and facilitators for the uptake of the HPV vaccine. For intervention studies, records will be included if they involve the evaluation of a digital/non-digital interventions to promote the uptake of the vaccine. The interventions that evaluate outcomes related to both distal and proximal social determinants of HPV vaccine uptake will be considered.

### Context

The concept of interest will include studies conducted in SSA across all healthcare levels from the primary care-level health facilities to the referral-level health facilities since 2006, when the first HPV vaccine was licensed^37^. Non-healthcare facilities such as schools will also be considered. Health equity is a key factor in health behaviour, so we will consider diverse studies conducted in different contexts including rural, urban, underserved, minoritized, and other populations.

### Type of evidence sources

The *determinants evidence* will be derived from peer-reviewed, non-peer reviewed and unpublished primary sources reporting qualitative, quantitative, and mixed-method studies of determinants of HPV vaccine uptake. *Interventions evidence* will be derived from peer-reviewed articles reporting quantitative studies of interventions effectiveness in promoting HPV vaccine uptake. Conference abstracts, reviews, editorials, letters to the editor and commentaries will be excluded.

## METHODS AND ANALYSIS

Since the publication of the seminal methodological framework for the conduct of scoping reviews^38^, followed by Levac and colleagues in 2010^39^, there has been a steady improvement with more recent developments of guidelines by the JBI Scoping Review Methodology Group^35,36,40^. The proposed scoping review will be conducted following the JBI methodology for scoping reviews^35,36^. The review will be reported according to the Preferred Reporting Items for Systematic Reviews and Meta-Analysis Extension for Scoping Reviews (PRISMA-ScR) checklist^41^.While the adoption of intersectionality approach in research has taken several dimensions including as a field of study, critical praxis, and as an analysis strategy, this review will apply the approach as an analytical framework^21^ to explore the interplay of multiple co-existing and interlocking social determinants that create inequities and inequalities of opportunity for HPV vaccine uptake. Thereafter, to integrate the *determinants evidence* with *interventions evidence*, a sequential explanatory design developed by the Evidence for Policy and Practice Information (EPPI) centre will be adopted^31,32^. This design examines the extent to which behaviour change interventions are aligned with reported contextual determinants of target behaviour. Notably, by adding complexity and complementarity lenses around complex intervention development, the overall methodological approach adopted for this review goes beyond a single-method review ^21,42,43^.

### Search strategy and information sources

The development of the search strategy and pilot testing was done in collaboration with a medical librarian. Moreover, the authors (P.K.) and (V.M.) will further collaborate with a medical librarian during the implementation of the search strategy. The search strategy aims to retrieve both published and non-peer-reviewed *(determinants evidence only)* literature related to the review questions. An initial preliminary search of PubMed was conducted for records on the topic. The keyword, free-texts contained in the titles and abstracts of relevant records, and the MeSH terms describing the records guided the development of a complete search syntax/strategy for PubMed/MEDLINE (supplementary file 1: Search strategy for PubMed/Medline). Thereafter, the search strategy was adapted for other included databases including LIVIVO, Google Scholar, BASE (Grey Literature), Preprints databases (e.g. OSF and MedRxiv), and African Journals Online (AJOL). Further search for relevant articles will be conducted on the reference lists of included records. Table 1 shows a sample search strategy for Medline via PubMed

**Table 1:**
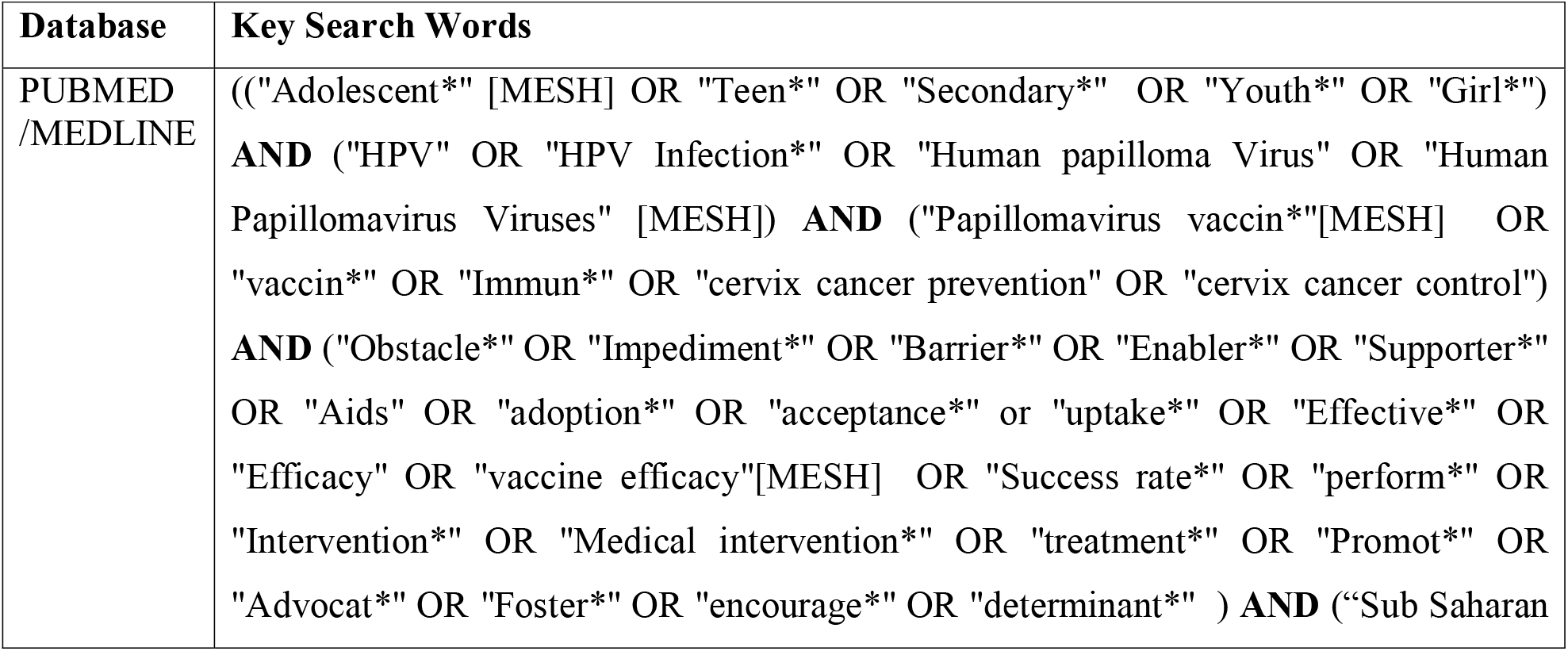
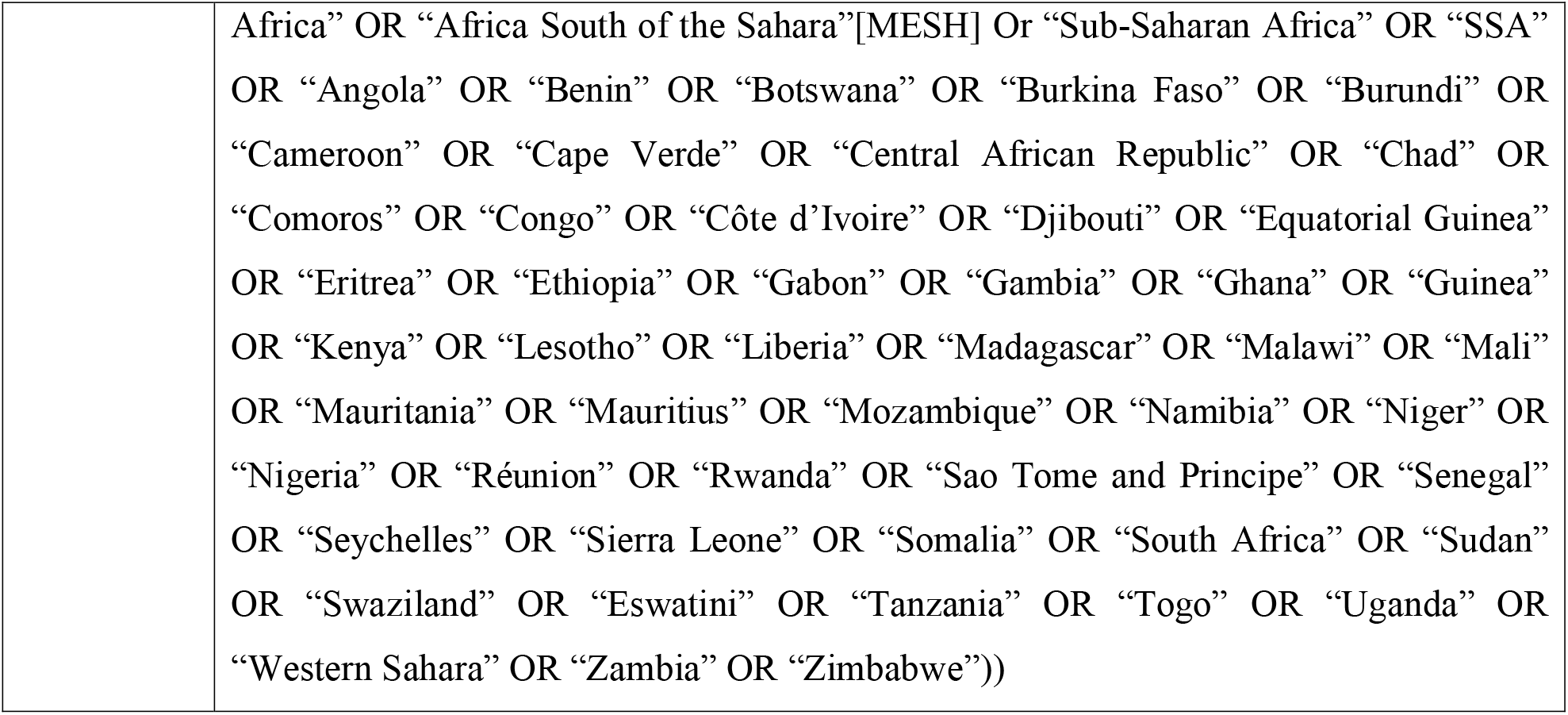
Search strategy for PubMed/Medline.

### Study selection

Following the implementation of the search strategy, retrieved records will be imported into EndNote citation management software. Thereafter, duplicates will be removed. The screening of the records will be conducted independently by the two reviewers in two phases beginning with titles and abstracts. Afterwards, full-text records will be screened based on the eligibility criteria. Any disagreements between the two reviewers about the eligibility of a study at any phase of the selection process will be resolved through consensus or consultations with a third independent party. Reasons for the exclusion of full-text records that do not meet the inclusion criteria will be recorded and reported within the review.

### Data charting/extraction

Just like in other stages of this scoping review, a team approach will be adopted during data extraction/charting^44^. While the specific data extraction items will be guided by the review questions, the overall process will be governed by the recently developed guideline around the data charting/extraction phase of scoping reviews^45^. In line with the sequential explanatory design underpinning this review, data will be extracted from the determinants studies and interventions studies. Two purposely developed data charting forms will be developed and independently pilot-tested by the two reviewers with any changes on the forms made collaboratively. Subsequently, changes to the forms will be made iteratively throughout the data extraction process as deemed necessary. The two reviewers will independently extract data from 50% of the included records, after which each of them will verify each other’s data extraction to ensure accuracy and completeness^46^. Any disagreements will be resolved through a consensus. For both determinants and interventions studies, data items to be extracted include citation, country, setting, type of study, methods, and participants. Data extraction items specific to determinants will be informed by the recently published WHO Operational Framework for Monitoring Social Determinants of Health Equity^47,48^. However, this framework will be used flexibly in consideration of the contextual embeddedness of social determinants of HPV vaccine uptake. For intervention studies, additional items to be extracted include aims, intervention content, duration, nature (digital or non-digital), complexity, and outcomes

### Data analysis and presentation

While cognizant of previous authors’ views that analysis in scoping reviews should be strictly descriptive^45^, the analysis adopted in this review will be informed by the review questions as well as the study design (sequential explanatory)^31,32^ underpinning this study. First, for determinants studies, thematic analysis developed by Thomas and Harden will be performed^49^. Furthermore, adoption of an intersectional analytic lens will enable multi-level analysis to expose intersections of determinants across and within socioecological levels that create inequalities of opportunity for HPV vaccine uptake. For intervention studies, regardless of the homogeneity or heterogeneity of studies, a narrative analysis will be performed. Lastly, a cross-study analysis will be conducted based on the Evidence for Policy and Practice Information (EPPI) centre approach for combining *determinants evidence* and *interventions evidence*^*32*^. This will compare the extent to which included interventions are sensitive to the participants’ views on determinants of the HPV vaccine uptake. Data will be presented in tables and matrices.

### Patient and public involvement

The research team plans to engage local adolescents, parents, and teachers to comment on the findings on social determinants of HPV uptake as well as the interventions to promote the uptake. Particularly, they will be invited to comment on the appropriateness of the current interventions in influencing the determinants for optimized HPV vaccine uptake.

## ETHICS AND DISSEMINATION

No ethical approval will be required for this study because it will be based on data collected from publicly available documents. However, all included studies will be assessed for adherence to ethical requirements. If any ethical inadequacies are found in the included studies, they will be acknowledged. For documents that don’t adequately report ethical considerations, authors will be contacted to obtain additional information. We will engage all relevant stakeholders including parents, adolescents, healthcare professionals, policymakers, healthcare administrators, cervical cancer survivors, and community-based organisations, to co-design strategies for the dissemination of review results. Particularly, the results will be shared with stakeholders directly involved with the uptake of the HPV vaccine, including clinicians, adolescents and their parents, healthcare administrators, and policymakers. Furthermore, the review will be written up as a journal article and submitted to a peer-reviewed journal.

### Strengths and limitations of this study

- This will be the first scoping review for the study of HPV vaccine uptake to adopt an intersectional lens as an analytical framework
- Inclusion of grey literature in the search strategy will broaden the number of papers retrieved
- The review covers a wide period of time from 2006 when the first HPV vaccine was licensed up to 2024.
- The adoption of mixed method review (sequential explanatory design) will enable integration of complementary evidence on HPV vaccine uptake
- The exclusion of non-English papers may narrow the scope of papers included in the review.

## Data Availability

All data produced in the present work are contained in the manuscript

## Authors’ contributions

PNK and VNM have all made substantial contributions to the conception or design of the manuscript and the acquisition of data. They have further made substantial contributions in the drafting and revising of the manuscript for important intellectual content, have had final approval of the version to be published and agreed to be accountable for all aspects of the work in ensuring that questions related to the accuracy or integrity of any part of the work are appropriately investigated and resolved.

## Disclaimer

The views expressed in the submitted article are those of the authors and not an official position of the authors’ institution

